# Development and performance evaluation of a low-cost in-house rRT-PCR assay in Ecuador for the detection of SARS-CoV-2

**DOI:** 10.1101/2021.08.03.21260966

**Authors:** Marco A. Salinas, Diana F. Aguirre, David I. De la Torre, Jorge Pérez-Galarza, Ronny J. Pibaque, Paul A. Beltran, Tatiana J. Veloz, Lucy Y. Baldeón

## Abstract

**Antecedents:** Ecuador has had the greatest fatality rate from Coronavirus (COVID-19) in South America during the SARS-CoV-2 pandemic. To control the pandemic, it is necessary to test as much population as possible to prevent the spread of the SARS-CoV-2 infection. For the Ecuadorian population, accessing a PCR test is challenging, since commercial screening kits tend to be expensive. Objective: the objective of this study was to develop an in-house duplex rRT-PCR protocol for the detection of SARS-CoV-2 that contributes to the screening while keeping quality and low testing costs. Results: An in-house duplex rRT-PCR protocol based on the viral envelope (E) gene target of SARS-CoV-2 and a human ribonuclease P gene (RP) as an internal control is reported. The protocol was optimized to obtain primers E with an efficiency of up to 94.45% and detection of 100% of SARS-CoV-2 up to 15 copies per uL. The clinical performance was determined by a sensibility of 93.8% and specificity of 98.3%. Conclusion: we developed, standardized, and validated a low-cost, sensitive in-house duplex rRT-PCR assay that may be utilized in low-income countries.

## 1. Introduction

On March 11, 2020, the World Health Organization (WHO) designated coronavirus disease 2019 (COVID-19) outbreak a pandemic and an international public health emergency [1]. COVID-19 disease is caused by a novel coronavirus, which is highly infectious and can induce severe acute respiratory syndrome (SARS-CoV2). Until June 2021, the number of cases reported is more than 180 million infections and more than 3 million deaths around the world [2]. The World Health Organization (WHO) recorded 360 thousand confirmed illnesses and over 18 thousand fatalities in Ecuador.

Ecuador has the highest case fatality rate in the area (4.9 percent) [3]. Furthermore, the country’s death rate increased by 24% in the first trimester of 2021 (mortality comparison between December 2019 and March 2020 vs. December 2020 and March 2021) [4].

The existence of asymptomatic persons confuses the precise number of infected patients, which is a possible danger that stays latent for the disease’s spread. As a result, the rate of SARS-CoV-2 infection may be greater than previously thought. [5]. This scenario emphasizes population-wide testing and, as a result, the availability of specialized tests for early viral identification and the implementation of pandemic control epidemiological methods. [6]. Ecuador is one of the countries in South America with the lowest number of tests per capita (0.2 per 1000 persons each day) [7].

The gold standard for SARS-CoV-2 detection is a viral nucleic acid detection technique based on real-time reverse transcriptase-PCR (rRT-PCR). Commercial kits based on rRT-PCR have been in great demand across the world due to the technique’s excellent sensitivity and specificity in accurately detecting the virus [8]. The US Food and Drug Administration (FDA) has approved various rRT-PCR assays targeting viral genes such as nucleocapsid (N), envelope (E), and RNA-dependent RNA polymerase (RNA-dependent RNA polymerase) (RdRp) [9]. However, numerous tests have shown that the E gene is somewhat more sensitive than other genetic targets, and the Pan American Health Organization (PAHO) has advised that the E gene be utilized exclusively for viral population screening in the Americas during this emergency [10].

Because of the COVID-19 pandemic, there has been an increase in the global demand for rRT-PCR commercial kits for detecting SARS-CoV-2. The scarcity of kits is worse in low-income countries like Ecuador, where the cost of these kits makes them inaccessible to the government and the general population. Many laboratories have created in-house assays with excellent sensitivity and specificity to tackle this challenge by emphasizing test cost reduction. In addition, to minimize the transmission of the virus by asymptomatic persons, the majority of the population must be screened. Our goal was to develop an in-house duplex rRT-PCR test that would identify SARS-CoV-2 in human respiratory samples using the E gene.

## 2. Materials and Methods

### 2.1. Primers and probes

For the duplex standardization experiment, two sets of primer pairs and probes for the viral E gene and human ribonuclease P (RP) (internal control) were employed. A multiplex assay was also attempted using primers targeting RdRp gene. E and RdRp primers/probes were obtained from the study published by Charité-Universitatsmedizin Berlin Institute of Virology [11]. RP primer/probe was obtained from the United States of America Centre for Disease Control and Prevention (CDC) [12]. The probes were labelled with FAM, ROX, and VIC for E, RdRp, and RP, respectively (see Table 1). All primers and probes were purchased from Eurofins Scientific (Kentucky, USA).

**Table 1.**
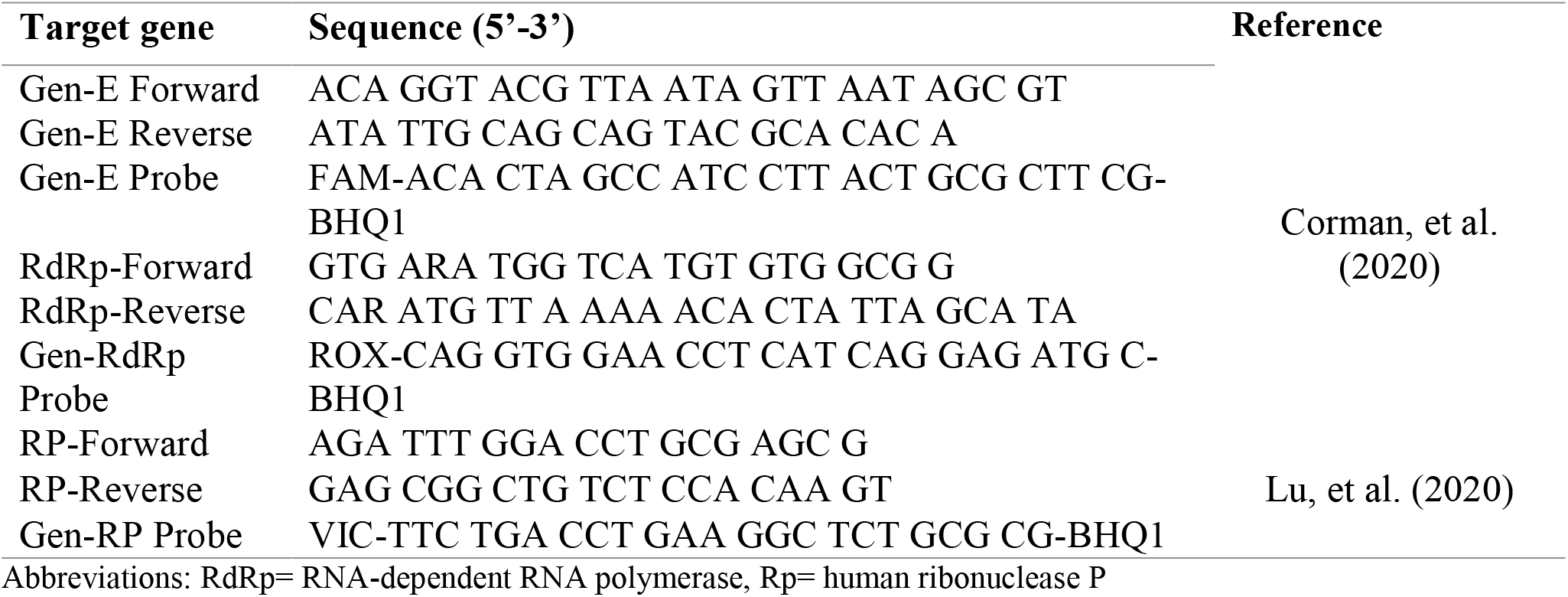
Primers and Probes used in Duplex rRT-PCR.

### 2.2. rRT-PCR reaction optimization

To develop the assay, a SARS-CoV-2 RNA source (RNA standardization solution) with a known concentration (3.81 x 105 viral copies per uL) was used. To optimize the final reaction setup, simplex assays were done. Three sets of primers/probes were used in different concentrations. The thermocycler program was set up using temperature gradient at 56, 58, and 60 °C. Amplification products were observed by electrophoresis (2% agarose gel) to verify the specificity of the primers. With 5 and 3.3 uL of RNA standardization solution, respectively, a final volume of 20 and 10 uL was tested. The master mix was prepared with the GoTaq Probe 1-step RT-qPCR System reagent (Promega, USA). The QuantStudio 5 Real-Time PCR equipment (Applied Biosystems, USA) and BioRad CFX96 (BioRad Laboratories, USA) were used to perform the experiments.

### 2.3. In-house duplex and multiplex rRT-PCR develop

5 uL of 1X GoTaq Probe qPCR Master Mix, 0.2 uL of 50X GoScript RT Mix 1-Step RT-qPCR, 30 nM CRX reference dye (only for QuantStudio 5), 400/200 nM primers/probe E gene, 200/100 nM primers/probe RP gene, and 3.3 uL of RNA standardization solution were used to make an in-house duplex assay. Thermal cycling conditions were 45 °C for 15 min, 95 °C for 2 min, and 40 cycles of 95 °C for 15 secs and 60 °C for 30 secs. For the multiplex assay, 300/150 nM primers/probe RdRp were added to the duplex setup and RNA standardization solution was decreased to 2.5 uL.

### 2.4. Limit of detection (LoD) of duplex rRT-PCR

To assess the efficiency of E primers, successive dilutions of the RNA standardization solution were produced in the range of 512 to 16 copies per uL. Serial dilutions were reduced from 64 to 1 copy per uL for the LoD of the duplex test. Three distinct operators carried out each test independently, and the reactions were produced in triplicate.

### 2.5. Clinical evaluation and ethical approval

The in-house assay was created and tested at the Research Institute in Biomedicine of Central University of Ecuador (INBIOMED-UCE). The researchers utilized stored RNA from nasopharyngeal swab samples from COVID-19 suspects who visited INBIOMED in February 2021. The Ministry of Public Health of Ecuador ethically approved the in-house SARS-CoV-2 assay.

### 2.6. Agreement and Performance

The sample was calculated to achieve a 95 percent level of significance and an 8 percent inference error. Taking into consideration a 25% probability of positive. As a result, the sample’s computed N was 112, which was raised by 10% to account for probable losses, yielding 124 samples. There were 64 COVID-19 positive samples and 60 COVID-19 negative samples in the study. To evaluate the agreement/performance, we compared the in-house duplex rRT-PCR results against the commercial LightMix SarbecoV E-gene plus EAV control kit and LightMix Modular SARS-CoV-2 (COVID-19) RdRp (TibMolBiol. Germany). For the LightMix E/RdRp kit, the samples were diagnosed as positive for SARS-CoV-2 when at least one viral gene had a valid CT value [10].

### 2.7. Viral RNA Extraction and in-house duplex rRT-PCR

The Nucleic Acid Extraction kit (Magnetic bead technique) (Zybio, China) was used to extract RNA from 200 uL of the sample, with an elution volume of 50 uL, according to the manufacturer’s instructions. The RNA was kept at -80 °C until it was needed. An in-house duplex technique was used to accomplish the rRT-PCR. The reactions for the LightMix E/RdRp kit were carried out according to the manufacturer’s instructions. Bio-Rad CFX96 was used to run the samples.

### 2.8. Statistical analysis

We calculate the positive percent agreement (sensitivity), negative percent agreement (specificity), positive and negative predictive values, likelihood ratio, and Cohen’s Kappa to assess the duplex rRT-PCR test in clinical samples. SPSS software version 23 was used for the analysis (IBM).

## 3. Results

### 3.1. rRT-PCR optimization

Simplex rRT-PCR experiments were prepared to find the optimal primers and probes concentration. The amplicons of these assays were located at the expected positions for the fragment lengths which were 113 bp, 100 bp, and 50 bp for the E, RdRp, and RP respectively. For the RP set, a low variation in the Ct was observed when changing RP primer concentrations from 600 nM to 200 nM (with half of the concentration of its probe). For this reason, 200/100 nM primers/probe concentration was chosen for all following assays. In the case of E and RdRp sets, 400/200 nM and 300/150 nM, respectively, was chosen for the remaining experiments because this primers/probe concentration had one of the lowest Ct (E=18.40, RdRp=25.65). As for the annealing temperature, the lowest Ct for the E pair of primers was prioritized. The temperature was 60 °C (Data not shown).

### 3.2. In-house duplex and multiplex SARS-CoV-2 assays

Upon optimization of reaction conditions on the simplex set-up we performed a duplex protocol, which had more consistent results than the multiplex protocol. When the Ct value of a target changed in more than one of the E and RP sets of primers/probes (Duplex) and E, RdRp, and RP (Multiplex) primers/probes, the reactions were declared invalid. E target had Ct values consistent across all assays (Ct 21.77 simplex, 21.83 duplex, 22.13 multiplex). RP readout, which was used to measure genetic material quality, exhibited similar Ct variation to the E gene, with no more than 0.5. (23.69 duplex, 24.19 multiplex). RdRp profiles were found to be less reproducible than the other two targets, although their Ct values were within a respectable margin of error between replicates (no more than 0.5) (see Figure 1). At low quantities of viral RNA (15 viral copies/reaction), the RdRp target showed weak or no amplification curves in further multiplex tests. With a final volume of 10 uL, all reactions were carried out. Reducing the reaction volume from 20 uL to 10 uL, and therefore the volume of RNA standardization solution from 5 uL to 3.3 uL, yielded the same findings without interfering with the amplification of E and RP targets. QuantStudio 5 and BioRad CFX96 were both compatible with the duplex test.

**Figure 1.**
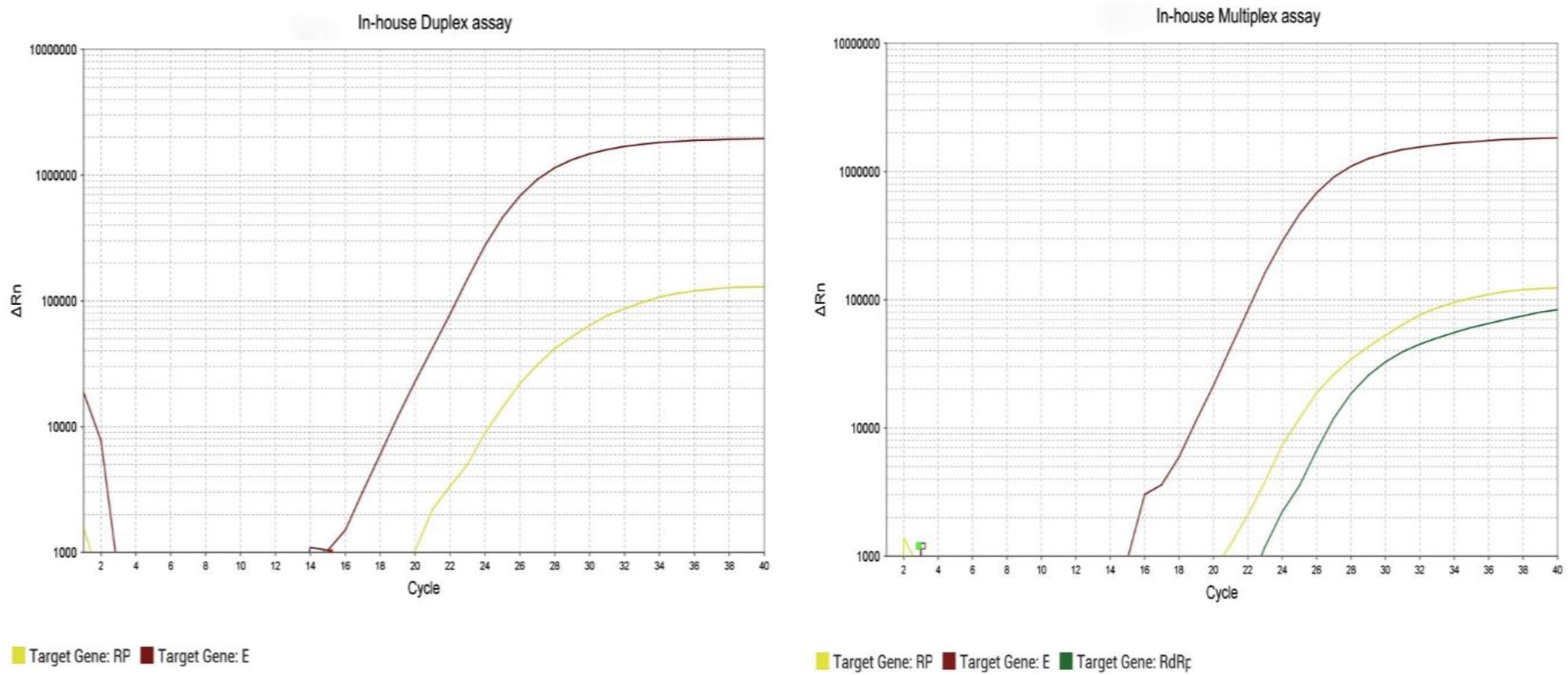
Amplification plots for duplex and multiplex protocols. Plots were obtained from a QuantStudio 5 Real-Time PCR system with detections of the target genes (E, RP, and RdRp).

### 3.3. LoD of the in-house duplex assay

Serial dilutions of the RNA standardization solution (64, 32, 15, 10, 5, 2, 1 copies/uL) were prepared for LoD evaluation. 15 copies/uL (49.5 copies/reaction) with a mean Ct value of 35.99 (CI95 percent 35.33 – 36.66) was the lowest concentration at which all nine replicates (100%) showed amplification curves. (see Table 2). Moreover, the primer efficiency was 94.45%, with an R2 of 98.1 percent. (see Figure 2).

**Table 2.** Confirmatory LoD Testing Results.

| Target level<br>(copies/uL) | N° Valid tested<br>replicates | SARS-CoV-2 E Gene Positive |  |  |
| --- | --- | --- | --- | --- |
|  |  | n* | Mean Ct | Detection rate |
| 64 | 9 | 9 | 34.05 | 100% |
| 32 | 9 | 9 | 35.25 | 100% |
| 15 | 9 | 9 | 35.99 | 100% |
| 10 | 9 | 5 | 35.74 | 55.56% |
| 5 | 9 | 6 | 37.23 | 66.67% |
| 2 | 9 | 4 | 35.60 | 44.44% |
| 1 | 9 | 2 | 35.89 | 22.22% |

\* This column contains only valid replicates

**Figure 2.**
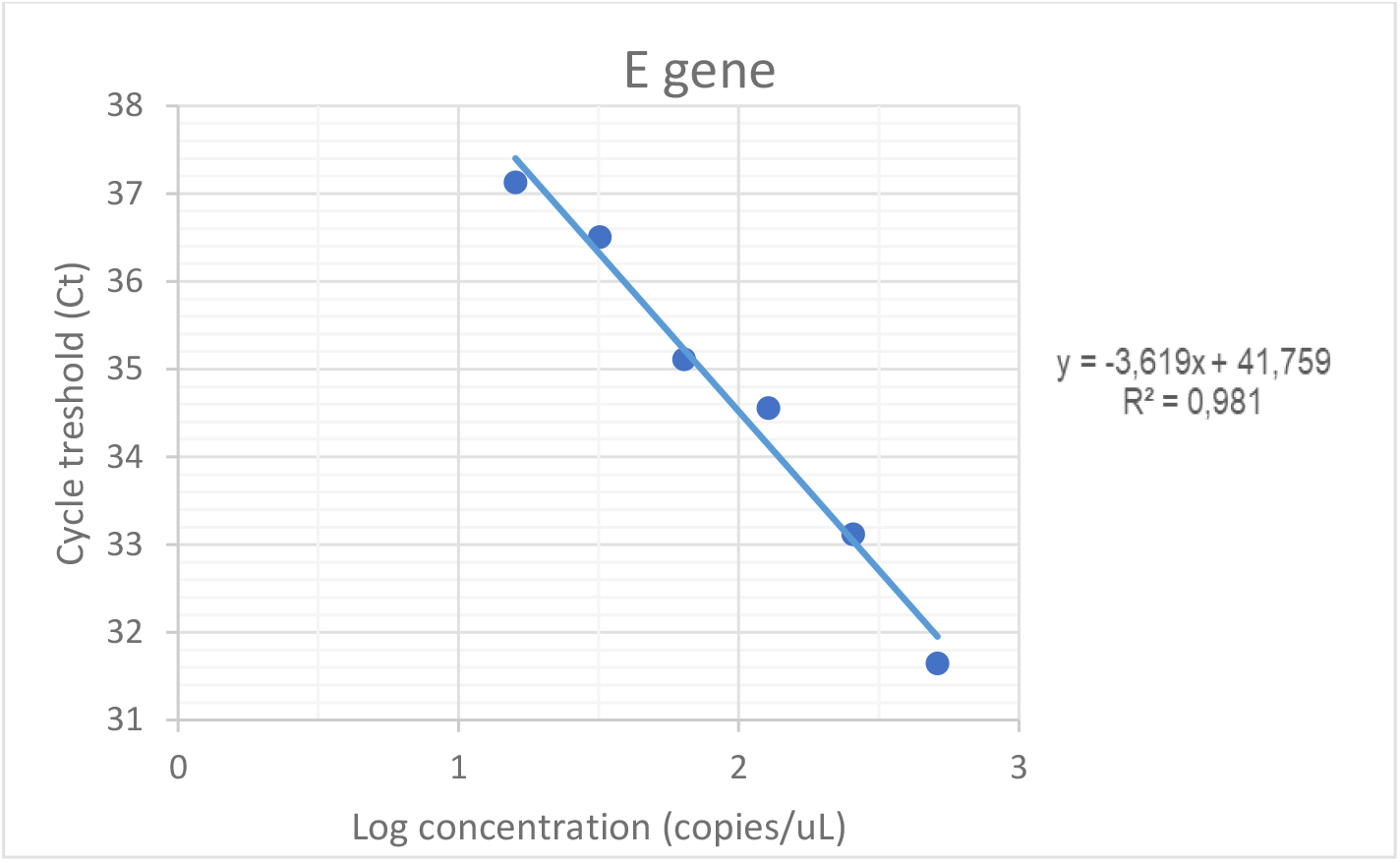
Standard curve plot of E gene from 2-fold serial dilutions of RNA standardization solution (512, 256, 128, 64, 32, 16 copies/uL). Primers’ efficiency = 94.45 %, slope -3.619 and R2 = 0.981.

### 3.4. Clinical Evaluation

For clinical comparison, we used the commercial LightMix SarbecoV E-gene + EAV control kit, which identified 64 positives and 60 negatives in nasopharyngeal samples. Sixty-four out of sixty-four samples were found to be positive using an in-house duplex method. The in-house duplex procedure found 60 out of 64 samples to be positive (see Table S1 in the Supplementary Material). Four samples were missed by the duplex test, resulting in 93.8 % sensitivity, 98.3 % specificity, a Likelihood ratio of 56.25, and a Cohen’s K of 0.92 (see Table 3).

**Table 3.** Clinical performance indicators of concordance of E target with in-house duplex rRT-PCR vs. LightMix E/RdRp kit.

|  | Sensitivity<br>(%) | Specificity<br>(%) | PPV | NPV | LR | Kappa |
| --- | --- | --- | --- | --- | --- | --- |
|  | (CI95%) | (CI95%) |  |  |  |  |
| In-house duplex<br>rRT-PCR | 93.8<br>(84.8-98.3) | 98.3<br>(91.1-100) | 98.4<br>(91.2-100) | 93.7<br>(84.5-98.2) | 56.25 | 0.92 |

Abbreviations: CI95%= Confidence Interval, PPV = Positive Predictive Value, NPV = Negative Predictive Value, LR = Likelihood Ratio, Kappa = Cohen's Kappa Index

## 4. Discussion

Ecuador is one of the South American countries worst hit by the COVID-19 pandemic. Molecular diagnostics using rRT-PCR is the best approach to detect SARS-CoV-2 virus to restrict its spread until we achieve herd immunity in the nation. An improved and reproducible rRT-PCR methodology for the diagnosis of COVID-19 using a viral gene (E) and a human gene (Rp) in a simple reaction was designed in this work, using primers built according to recognized international guidelines [11,12]. As a result, 113-bp bands for the E gene and 55-bp bands for the Rp gene were produced without nonspecific products such as dimers or overlapping sequences that might cause false positives during the amplification process [13].

The sets of primers/probes for the detection of the E and RdRp genes employed to standardize the technique have a reasonably low nonselective mutation rate and have been reported in similar researches [14,15]. Following the guidelines of the US Centers for Disease Control and Prevention (CDC), the Rp gene was utilized as an internal control of RNA extraction [16]. The E gene showed little variability in duplex and multiplex reactions, according to the amplification graph (Figure 1). In contrast, using the RdRp gene, repeatability and effectiveness were dramatically reduced at low viral doses. In accordance with findings from numerous studies, which suggest that the RdRp gene set of primers/probe is less sensitive than the E gene [17,18]. For this reason, the E gene duplex combination was chosen. In contrast to Corman et al., who reported a LoD of 3.9 copies/reaction (95% CI: 2.8–9.8) [11]., we detected 49.5 copies/reaction for the E gene with a probability of 100 % detection (Table 2). The type of reagents and equipment used during the test might explain the variation in LoD [19-21].

The clinical assessment of the kit revealed that it has a diagnostic sensitivity of 93.8 % and a specificity of 98.3 %, implying a 6.2 % false negative rate and a 1.7 % false positive rate. In the assessed population, the degree of agreement of the findings between in-house rRT-PCR and the commercial LightMix SarbecoV gene E kit is strong (Kappa index = 0.92). Furthermore, our kit has a positive predictive value of 98.4%, indicating that in a positive test, this percentage of persons actually has the condition. The 93.7 % negative predictive value indicates that 93.7 percent of those who get a negative screening test do not have the disease. Meanwhile, the odds of exposure among case-patients are 56 times greater than the odds of exposure among controls.

The limitation of the in-house duplex test is that it can only identify one viral gene (E) and one internal control gene (Rp). However, the kappa index (0.92) shows that our protocol is comparable to commercial kits (LightMix E / RdRp) that screen two or more viral genes. Furthermore, because the E gene is unique to all Sarbecoviruses and because SARS-CoV-2 is the only member of the family now circulating in humans, the WHO has recommended that the E gene be prioritized as a target. In this approach, a single viral genetic target suffices for case confirmation in the lab.

## 5. Conclusions

In conclusion, we designed, standardized, and validated an in-house duplex rRT-PCR assay that can detect SARS-CoV-2 virus presence up to 15 copies/uL. This approach can assist our country enhance its capacity to screen both symptomatic and asymptomatic carriers by making SARS-CoV-2 rRT-PCR more accessible.

## Data Availability

The Tabular data set used to support the findings of this study have been deposited in the 4TU.RESEARCHDATA repository.

https://figshare.com/s/1dccb42f2189fbd42a65

## 6. Ethical Statement

This study was approved by Health Intelligence Directorate of the Ministry of Public Health of Ecuador through resolution No MSP-CGDES-2020-0259-O.

## 7. Data Availability

The [TABULAR DATASET] data used to support the findings of this study have been deposited in the [4TU.RESEARCHDATA] repository (https://figshare.com/s/1dccb42f2189fbd42a65).

## 8. Conflict of interest

The authors declare that there is no conflict of interest regarding the publication of this paper.

## 9. Funding

This work was supported by the Académie De Recherche Et D’Enseignement Supérieur – Central University of Ecuador (ARES-UCE) and Empresa Pública de Bienes y Servicios -Central University of Ecuador (EP-UCE).

## 10. Acknowledgment

We are extremely grateful to Central University of Ecuador, for his support in managing the needed funds for the development of this in-house duplex rRT-PCR protocol that would have a great positive impact in our society.

## 11. Supplementary Materials

The Table S1 used to support the findings of this study has been deposited in the [4TU.RESEARCHDATA] repository (https://figshare.com/s/1dccb42f2189fbd42a65).

## Notes

### Competing Interest Statement

The authors have declared no competing interest.

