## Supplementary material for "Development and performance evaluation of a low-cost in-house rRT-PCR assay in Ecuador for the detection of SARS-CoV-2": Suplementary Table S1

Table S1. Ct values of 124 clinical samples done with in-house duplex assay and LightMix E/RdRp commercial kit.

|  | In house Duplex assay | | LightMix E/RdRp kit | | |
| --- | --- | --- | --- | --- | --- |
| No | E gene | Rp gene | E gene | IC gene | RdRp gene |
| 1 | 31.79 | 26.20 | 30.70 | 33.48 | 29.35 |
| 2 | 23.28 | 24.44 | 22.17 | 25.18 | 29.06 |
| 3 | 23.75 | 24.52 | 22.83 | 26.44 | 29.07 |
| 4 | 26.54 | 25.93 | 25.08 | 28.58 | 29.20 |
| 5 | 20.24 | 22.73 | 18.84 | 22.76 | 29.03 |
| 6 | 23.28 | 25.17 | 22.13 | 25.10 | 29.11 |
| 7 | 24.44 | 25.52 | 23.12 | 26.26 | 29.12 |
| 8 | 22.23 | 24.11 | 21.37 | 24.59 | 29.18 |
| 9 | 26.27 | 25.58 | 25.24 | 29.44 | 29.28 |
| 10 | 28.02 | 25.88 | 26.28 | 30.49 | 29.26 |
| 11 | 23.16 | 25.31 | 22.12 | 25.38 | 29.18 |
| 12 | 35.08 | 27.25 | 34.79 | 39.54 | 29.33 |
| 13 | 22.51 | 23.92 | 21.11 | 24.80 | 29.16 |
| 14 | 17.07 | 22.54 | 21.38 | 24.39 | 29.07 |
| 15 | 19.16 | 21.71 | 16.97 | 20.99 | 29.20 |
| 16 | 26.02 | 24.19 | 24.81 | 29.48 | 29.13 |
| 17 | 29.45 | 25.10 | 28.50 | 31.75 | 29.08 |
| 18 | 29.72 | 25.79 | 28.02 | 31.96 | 29.07 |
| 19 | 29.40 | 26.16 | 31.83 | 38.84 | 29.28 |
| 20 | 18.46 | 21.25 | 17.38 | 21.77 | 29.39 |
| 21 | 26.16 | 25.14 | 25.23 | 28.90 | 29.25 |
| 22 | 21.15 | 23.53 | 20.44 | 24.23 | 29.30 |
| 23 | 29.34 | 25.77 | 28.27 | 32.14 | 29.25 |
| 24 | 22.03 | 24.63 | 21.37 | 25.39 | 29.19 |
| 25 | 18.82 | 21.91 | 17.77 | 23.01 | 29.16 |
| 26 | 23.48 | 24.22 | 22.92 | 27.28 | 29.03 |
| 27 | 34.55 | 24.45 | 33.91 | 37.32 | 29.34 |
| 28 | 25.43 | 26.48 | 23.85 | 27.57 | 29.17 |
| 29 | 20.34 | 20.34 | 20.62 | 24.33 | 28.95 |
| 30 | 28.58 | 28.58 | 29.87 | 32.41 | 28.32 |
| 31 | 33.08 | 33.08 | N/D | N/D | 28.58 |
| 32 | 27.32 | 27.32 | 30.49 | 33.54 | 28.42 |
| 33 | 29.89 | 29.89 | 29.13 | 32.91 | 29.85 |
| 34 | 19.41 | 19.41 | 33.81 | 36.65 | 28.86 |
| 35 | 30.79 | 30.79 | 31.21 | 33.93 | 28.60 |
| 36 | 18.85 | 18.85 | 18.88 | 20.46 | 28.32 |
| 37 | 16.40 | 16.40 | 15.56 | 18.23 | 28.16 |
| 38 | 20.10 | 20.10 | 19.10 | 20.07 | 28.16 |
| 39 | 17.07 | 17.07 | 15.31 | 17.38 | 28.15 |
| 40 | 25.51 | 25.51 | 24.97 | 26.77 | 28.31 |
| 41 | 18.28 | 18.28 | 18.05 | 19.37 | 28.40 |
| 42 | 31.07 | 31.07 | 30.22 | 32.27 | 28.48 |
| 43 | 18.41 | 18.41 | 31.44 | 33.50 | 28.58 |
| 44 | 15.14 | 15.14 | 15.67 | 17.44 | 28.31 |
| 45 | 27.03 | 27.03 | 25.65 | 27.18 | 28.06 |
| 46 | 14.46 | 14.46 | 15.11 | 19.31 | 28.34 |
| 47 | 18.74 | 18.74 | 30.15 | 31.20 | 28.42 |
| 48 | 21.12 | 21.12 | 22.28 | 26.30 | 28.14 |
| 49 | 20.24 | 20.24 | 19.38 | 20.68 | 28.07 |
| 50 | 20.91 | 20.91 | 29.80 | 34.24 | 28.35 |
| 51 | 17.02 | 17.02 | 15.95 | 16.95 | 28.06 |
| 52 | 38.45 | 38.45 | 34.99 | 37.02 | 28.25 |
| 53 | 30.41 | 30.41 | 19.14 | 34.61 | 28.18 |
| 54 | 26.62 | 26.62 | 17.60 | N/D | 28.50 |
| 55 | 22.27 | 22.27 | N/D | 24.66 | 28.97 |
| 56 | 19.24 | 19.24 | 19.86 | 20.40 | 28.39 |
| 57 | 17.84 | 17.84 | 26.76 | 20.78 | 28.33 |
| 58 | 27.38 | 27.38 | 31.52 | N/D | 28.34 |
| 59 | 18.78 | 18.78 | 34.61 | 22.22 | 28.49 |
| 60 | 26.51 | 26.51 | 22.18 | 28.19 | 28.37 |
| 61 | 27.03 | 27.03 | 21.29 | 23.09 | 28.19 |
| 62 | N/D | 26.33 | N/D | N/D | 29.36 |
| 63 | N/D | 26.15 | N/D | N/D | 29.41 |
| 64 | N/D | 25.79 | N/D | N/D | 29.25 |
| 65 | N/D | 21.75 | N/D | N/D | 29.20 |
| 66 | N/D | 26.64 | N/D | N/D | 29.23 |
| 67 | N/D | 27.34 | N/D | N/D | 29.33 |
| 68 | N/D | 26.18 | 44.39 | N/D | 29.29 |
| 69 | N/D | 27.47 | N/D | N/D | 29.32 |
| 70 | N/D | 27.35 | 40.20 | 40.63 | 29.06 |
| 71 | N/D | 26.43 | N/D | N/D | 29.33 |
| 72 | N/D | 26.08 | N/D | N/D | 29.32 |
| 73 | N/D | 26.37 | 41.93 | N/D | 29.25 |
| 74 | N/D | 26.01 | 40.95 | N/D | 29.11 |
| 75 | N/D | 26.21 | 41.29 | N/D | 29.40 |
| 76 | N/D | 26.09 | 42.96 | N/D | 29.27 |
| 77 | N/D | 25.97 | N/D | N/D | 29.28 |
| 78 | N/D | 26.72 | N/D | N/D | 29.34 |
| 79 | N/D | 27.31 | 39.24 | N/D | 29.41 |
| 80 | N/D | 26.73 | 42.61 | N/D | 29.57 |
| 81 | N/D | 28.66 | N/D | N/D | 32.16 |
| 82 | N/D | 26.99 | 41.82 | N/D | 29.46 |
| 83 | N/D | 26.23 | N/D | N/D | 29.88 |
| 84 | N/D | 26.12 | 41.22 | N/D | 29.22 |
| 85 | N/D | 23.63 | 44.32 | N/D | 29.30 |
| 86 | N/D | 24.55 | N/D | N/D | 29.64 |
| 87 | N/D | 26.34 | N/D | N/D | 29.49 |
| 88 | N/D | 24.70 | N/D | N/D | 29.25 |
| 89 | N/D | 24.98 | 38.35 | N/D | 30.20 |
| 90 | N/D | 24.36 | 37.37 | N/D | 29.24 |
| 91 | N/D | 24.73 | 43.68 | N/D | 29.22 |
| 92 | N/D | 23.25 | N/D | N/D | 28.37 |
| 93 | N/D | 24.54 | N/D | N/D | 28.35 |
| 94 | N/D | 24.59 | N/D | N/D | 28.35 |
| 95 | N/D | 21.08 | N/D | N/D | 28.10 |
| 96 | N/D | 23.40 | N/D | N/D | 28.20 |
| 97 | N/D | 24.90 | N/D | N/D | 28.23 |
| 98 | N/D | 24.21 | N/D | N/D | 28.20 |
| 99 | N/D | 24.69 | N/D | 38.20 | 28.10 |
| 100 | N/D | 25.37 | N/D | N/D | 28.23 |
| 101 | N/D | 24.44 | N/D | N/D | 28.56 |
| 102 | N/D | 24.65 | 36.66 | 38.16 | 28.34 |
| 103 | N/D | 24.85 | N/D | N/D | 28.66 |
| 104 | N/D | 24.60 | 37.99 | N/D | 29.78 |
| 105 | N/D | 25.02 | 39.32 | N/D | 29.50 |
| 106 | N/D | 26.56 | N/D | N/D | 28.36 |
| 107 | N/D | 25.68 | N/D | N/D | 28.29 |
| 108 | N/D | 26.38 | N/D | N/D | 28.40 |
| 109 | N/D | 26.42 | N/D | N/D | 28.28 |
| 110 | N/D | 26.30 | N/D | N/D | 28.31 |
| 111 | N/D | 28.60 | N/D | N/D | 28.38 |
| 112 | N/D | 26.94 | N/D | N/D | 28.32 |
| 113 | N/D | 27.30 | N/D | N/D | 28.22 |
| 114 | N/D | 28.67 | N/D | N/D | 28.38 |
| 115 | N/D | 25.09 | N/D | 39.17 | 28.28 |
| 116 | N/D | 25.65 | N/D | N/D | 28.39 |
| 117 | N/D | 26.64 | 41.83 | 39.20 | 28.37 |
| 118 | N/D | 27.82 | N/D | N/D | 28.42 |
| 119 | N/D | 26.65 | N/D | N/D | 28.28 |
| 120 | N/D | 25.99 | N/D | N/D | 28.31 |
| 121 | N/D | 26.52 | N/D | N/D | 28.21 |
| 122 | N/D | 25.08 | N/D | N/D | 28.30 |
| 123 | N/D | 26.14 | N/D | N/D | 28.34 |
| 124 | N/D | 25.70 | N/D | N/D | 28.32 |

Abbreviations: IC = Internal control, RdRp= RNA-dependent RNA polymerase, Rp= human ribonuclease P.
